# Dynamic Risk Stratification for Short-Term Deterioration After Acute Stroke: Incremental Value of Radiology-Report-Derived Neuroimaging Phenotypes Beyond Dynamic ICU States — a retrospective cohort study with temporal validation

**DOI:** 10.64898/2026.10.08.26365059

**Authors:** Ping Lei, Yanyi Xu, Yuxia Zhang, Nicolas Ray

## Abstract

**Background:** Prediction models after stroke increasingly add neuroimaging information to bedside data. Whether findings documented in radiology reports improve short-term risk stratification beyond a patient’s evolving physiological state is unknown.

**Methods:** We studied 4,827 adults with acute ischaemic stroke, intracerebral haemorrhage or subarachnoid haemorrhage in MIMIC-IV (2008–2019). At eight landmarks from 6 to 48 hours after ICU admission, models ranked patients by their risk of deterioration or death within 24 hours. The baseline model combined demographic and clinical characteristics, a previously validated dynamic ICU state representation, and the physiology, treatment and imaging status at each landmark. A locally run language model extracted imaging phenotypes from 6,992 head CT and MRI reports; three phenotypes and four location categories met a pre-specified reliability criterion for automated extraction in independent validation and entered the model. The primary outcome was the difference in the proportion of events captured among the 10% of patients ranked highest at each landmark, in temporal validation (2017–2019; 8,923 patient-landmarks, 666 events).

**Results:** Among the highest-ranked 10%, adding imaging phenotypes captured no additional event (348 of 666 with and without them): a difference of 0.00 percentage points (95% CI −2.20 to +2.01). Discrimination (AUROC 0.855 vs 0.849), calibration and net benefit were similar. No prespecified sensitivity analysis, stroke subtype or availability-adjusted analysis showed statistically supported incremental capture. In a post hoc redundancy analysis, imaging phenotypes improved capture when added to demographic and clinical characteristics alone (+12.91 points, 7.54 to 17.81), but this gain was no longer evident once the dynamic state representation was included.

**Conclusions:** Neuroimaging phenotypes derived from routine radiology reports were associated with short-term deterioration, but we found no evidence that they added risk stratification value beyond the dynamic ICU state and contemporaneous physiology.

## Background

Patients admitted to intensive care after acute stroke can deteriorate quickly and unevenly. Haematoma expansion, malignant oedema with herniation, and respiratory failure requiring ventilation develop over hours to days, and a patient who is stable at one assessment may need airway support at the next [1–3]. Identifying, among the patients in a unit at a given moment, those most likely to deteriorate in the next day is a practical triage problem: it decides who is reviewed first and who is watched most closely.

Neuroimaging is a natural candidate to sharpen that judgement. It defines the lesion: lateral displacement of the brain tracks depressed consciousness [4], and intraventricular haemorrhage is a marker of severity and poor outcome after intracerebral haemorrhage [5]. Prediction models increasingly add imaging information, often extracted from radiology reports, to structured clinical data, and some report gains in discrimination [6,7]. These models have predicted from a single time point against baselines of admission data, and one drew on reports from the whole hospital stay rather than those available when a prediction would be made [6]; a recent intensive care model of respiratory failure after ischaemic stroke could not include imaging at all [8].

A strong dynamic baseline changes the question. We previously described four clinical states that patients move between during the first 72 hours of ICU care after stroke [9] and examined their transportability across 176 hospitals in an independent multicentre database [10]. If such a representation already reflects the clinical consequences of a lesion — reduced consciousness, loss of airway protection — then a report describing that lesion may add less to a ranking of who deteriorates next than it would to a static model.

We therefore asked whether adding radiology-report-derived neuroimaging phenotypes to a model that already knows a patient’s dynamic state, current physiology and whether they have been imaged improves the identification of patients who deteriorate or die within 24 hours, at repeated landmarks during the first two days of ICU care after stroke.

## Methods

### Study design

This was a retrospective prognostic study with repeated landmarks, reported following TRIPOD+AI [11]. The analysis protocol was fixed before the imaging extractor was validated and before any model containing imaging content was fitted; later amendments were recorded with dates and versions (Additional file 1). The study was not registered, and no patients or members of the public were involved in its design, conduct or reporting.

### Data source and participants

MIMIC-IV (version 3.1) contains de-identified records of patients admitted to the intensive care units of Beth Israel Deaconess Medical Center, Boston [12–14]; radiology reports come from MIMIC-IV-Note, version 2.2 [15]. Access was granted under the PhysioNet credentialed data use agreement. The data are de-identified, and the Institutional Review Board of Beth Israel Deaconess Medical Center waived informed consent for their collection and release (Declarations). The source cohort comprised adults admitted to intensive care with a primary diagnosis of acute ischaemic stroke (AIS), intracerebral haemorrhage (ICH) or subarachnoid haemorrhage (SAH), as defined previously [9,10]. Because radiology reports are available only until 2019, admissions were restricted to 2008–2019. Admissions from 2008–2016 formed the development cohort and those from 2017–2019 the temporal validation cohort; no patient contributed to both.

### Landmarks, dynamic state and outcome

ICU time was divided into consecutive 6-hour windows. Landmarks were placed at the end of each window from 6 to 48 hours after admission, eight in all. Predictors were assigned to each landmark using the timing rules inherited from the earlier study: radiology reports were considered available only after they had been stored, whereas laboratory values were assigned to windows by specimen collection time. On audit, 17.4% of the laboratory values used at a landmark were resulted after it (median delay among these 0.60 hours; 90th percentile 1.56 hours).

Clinical state was represented by the treatment-free version of the previously described four-state hidden Markov model, whose parameters were fixed before this study. That model and its preprocessing (imputation values and scaling) had been estimated in the earlier study on a random split of 2008–2022 admissions, which included 913 of the 1,318 temporal-validation patients (69.3%); no outcomes were used in estimating it. So that deterioration would not be defined by treatment itself, this version excludes the direct treatment variables — mechanical ventilation, renal replacement therapy, vasopressors and sedation — and is estimated from the 17 physiological and laboratory measures alone. The same model supplied the real-time state used for prediction and eligibility and the states used to determine the outcome; treatments entered the prediction models only as predictors.

Standard decoding assigns a state to each window using the whole sequence of observations, so later data influence earlier states; such states cannot be used for prediction. At each landmark we therefore used the filtered state: the posterior probability of each state given only the windows up to and including that landmark. The four probabilities entered the models as three log-ratios against a reference state, together with the posterior entropy.

A patient entered the risk set at a landmark if they were alive and still in the ICU and, according to the filtered state, not already in the adverse state — the treatment-free analogue of the previously described respiratory-support state. The outcome was entry into the adverse state, or death, within the next 24 hours, determined from the complete observed sequence. Patients discharged or transferred out of the ICU alive within 24 hours, without prior deterioration, were counted as having no event.

### Models

Models were built in steps, each adding one source of information:

- **Dynamic-state baseline (M0-state):** age, sex, Charlson comorbidity index, hypertension, diabetes, atrial fibrillation, heart failure, chronic kidney disease, stroke subtype, landmark time, and the filtered state.
- **Full physiological baseline (M0-full):** adds 17 physiological and laboratory measures and four treatments (mechanical ventilation, renal replacement therapy, vasopressors, sedation).
- **Imaging-availability baseline (M0+A):** adds whether a head CT or MRI report was available at the landmark — whether the patient had been imaged, not what the scan showed. This 37-predictor model was the reference for the primary comparison.
- **Imaging-augmented model (M1-D):** adds imaging phenotypes where a report was available, and reduces to the imaging-availability baseline where it was not.
- **Availability-adjusted imaging-information model (M1-I):** estimates the information in imaging phenotypes among landmarks with a report, weighted for who was imaged.

All were L2-penalised logistic regressions on standardised predictors, fitted in the development cohort and applied unchanged to the validation cohort.

### Imaging phenotypes

#### Reports

Head CT and MRI reports from the index admission that were stored in the record by the 48-hour landmark formed the analysis population, including scans performed in the emergency department before admission. A report counted as available at a landmark from the time it was stored, not the time of the scan. Reports stored more than 72 hours before ICU admission were excluded as likely to predate the stroke.

#### Extraction

A large language model (Qwen2.5, 7 billion parameters, 4-bit quantised) run on local hardware through Ollama extracted findings from each report, so that no report text left the study computer [16]. Output was deterministic and constrained to a fixed ontology: each phenotype was recorded as present, absent, uncertain or not assessable, and lesion location as lists of categories. Rule-based checks verified the output against the report text. The extractor was developed on 50 reports and then fixed (Additional file 1).

#### Validation

The fixed extractor was compared with blinded manual annotation of 200 reports drawn at random from the analysis population, one per patient, stratified by stroke subtype and excluding the development reports and their patients. Annotation was performed by a single trained annotator following a written guideline. The validation therefore measures agreement with one annotator, not with a multi-expert consensus.

#### Reliability criterion

A phenotype or location category entered the primary model only if Cohen’s κ was at least 0.60, with at least 10 annotator-positive reports. The criterion was intended to decide whether automated extraction was reliable enough to enter the primary predictive model, not as a claim of clinical interchangeability with expert annotation. Findings below it were analysed only in sensitivity analysis, and the extractor was not modified after validation.

#### Use at landmarks

Evolving findings (haemorrhage, intraventricular haemorrhage, midline shift and haemorrhage compartment) were taken from the most recent eligible report; lesion location (vascular territory and anatomical region) was counted if reported in any eligible report. Present was coded as positive and absent as negative; uncertain and not assessable were treated as unknown rather than negative. Each finding entered the model as indicators for positive and for unknown, with negative as reference, so that landmarks without a report carried unknown for every finding. A report that could not be processed was treated as unavailable.

### Imaging availability and inverse probability weighting

Whether and when a patient is imaged reflects clinical judgement. The imaging-augmented model addresses deployment, in which only the images actually obtained are available, and was therefore not adjusted. For the availability-adjusted model, the probability of having an eligible report was estimated from the same pre-landmark predictors, and imaged landmarks were weighted by stabilised inverse probability weights truncated at the 1st and 99th percentiles, with common support assessed at each landmark.

### Statistical analysis

#### Primary comparison

The imaging-augmented model was compared with the imaging-availability baseline in the validation cohort. At each landmark, patients were ranked by predicted risk and the highest-ranked 10% were flagged. Flagged patients were pooled across landmarks, and the outcome was the difference in the proportion of all 24-hour events among them (event capture). Ranking within landmarks reflects the clinical question of which patients currently in the unit to prioritise; pooling provided the necessary precision, as no single landmark had more than 104 validation events. Confidence intervals came from 2,000 bootstrap resamples of patients, preserving the clustering of landmarks within patients.

#### Conclusion rule

If the 95% confidence interval lay entirely above zero, imaging phenotypes would be considered to add statistically supported value; otherwise, no incremental value would be concluded, without substitution by secondary results. A difference of 5 percentage points — at the 24-hour landmark, about four additional deteriorating patients among 117 flagged — was specified as a scale for clinical interpretation, not as a significance, equivalence or non-inferiority threshold.

#### Secondary analyses

We report capture, positive predictive value and lift (event rate among flagged patients relative to the landmark’s overall rate) at the top 5%, 10% and 20%; the areas under the receiver operating characteristic (AUROC) and precision–recall curves; Brier score; calibration intercept and slope, before and after intercept-only recalibration; and decision curves for threshold probabilities of 1% to 30% [17]. Results were examined within stroke subtypes, and a subtype-by-imaging interaction was tested because every subtype had at least 280 development events, the prespecified minimum.

#### Sensitivity analyses

Sixteen prespecified sensitivity analyses varied the outcome definition and horizon, the baseline and its state representation, the handling of unknown and uncertain findings, the phenotype set, the imaging update rule, the model class and the availability weighting (Table 4).

#### Post-audit analyses

After the primary result was known, an audit of the preprocessing inherited from the earlier study prompted two additional analyses that were not in the protocol and are not counted among the sensitivity analyses, each with its interpretation rule written down before it was run: the primary comparison restricted to validation patients not used to estimate the state model, and a rebuild in which laboratory values were assigned by result time and the laboratory predictors and filtered state recomputed, with everything else unchanged.

#### Repeated extraction with a larger model

After the primary result was known, the extraction was repeated with a larger locally run model (Qwen3.8, 27 billion parameters) [18], with the same prompt, rules and settings and without tuning, and the primary comparison was repeated on the same seven findings and on the findings that met the reliability criterion under the new extraction. The analysis and its interpretation rule were added to the protocol before the model processed any report (Additional file 1: Methods S6).

#### Post hoc redundancy analysis

After the primary result was known, we examined whether the prognostic information in imaging phenotypes overlapped with information already in the model. The same imaging variables were added to four nested baselines, in a fixed order: demographic and clinical characteristics with imaging availability; then the filtered state; then Glasgow Coma Scale eye and motor scores; then all physiology and treatment. The analysis and its interpretation rule were written down before it was run, but it was not part of the protocol.

The primary result was reproduced with independently written code (Additional file 1). Analyses used Python 3.9.6 with scikit-learn 1.6.1, pandas 2.3.3, NumPy 2.0.2 and SciPy 1.13.1; extraction ran on Ollama 0.13.1. The analysis code was written and run with the assistance of an AI coding assistant (Claude, Anthropic), which also helped draft and revise the manuscript text, and ChatGPT (OpenAI) was used for language editing of parts of the text. Neither tool had access to report text or patient-level data; the authors directed and reviewed all AI-assisted work and take responsibility for it.

## Results

### Participants

The analysis included 4,827 patients and 30,941 patient-landmarks (Table 1): 3,509 patients with 22,018 patient-landmarks and 2,190 events (9.9%) in development, and 1,318 patients with 8,923 patient-landmarks and 666 events (7.5%) in validation. Median age was 69 years (interquartile range 57–80); 50.0% were men, and 56.0% had AIS, 30.4% ICH and 13.3% SAH. Between 3,141 and 4,367 patients were at risk at each landmark, with event rates of 8.3% to 10.3%.

**Table 1.** Characteristics of patients at their first eligible landmark.

|  | All (n =<br>4,827) | Development 2008–2016<br>(n = 3,509) | Temporal validation 2017–2019<br>(n = 1,318) |
| --- | --- | --- | --- |
| Age, years, median (IQR) | 69 (57–80) | 69 (57–81) | 69 (57–80) |
| Male | 2,413 (50.0) | 1,767 (50.4) | 646 (49.0) |
| Stroke subtype |  |  |  |
| Acute ischaemic stroke | 2,703 (56.0) | 1,912 (54.5) | 791 (60.0) |
| Intracerebral haemorrhage | 1,465 (30.4) | 1,096 (31.2) | 369 (28.0) |
| Subarachnoid haemorrhage | 643 (13.3) | 488 (13.9) | 155 (11.8) |
| Both ICH and SAH | 16 (0.3) | 13 (0.4) | 3 (0.2) |
| Charlson comorbidity index,<br>median (IQR) | 3 (2–5) | 3 (1–5) | 3 (2–5) |
| Hypertension | 3,610 (74.8) | 2,617 (74.6) | 993 (75.3) |
| Diabetes | 1,297 (26.9) | 962 (27.4) | 335 (25.4) |
| Atrial fibrillation | 1,467 (30.4) | 1,073 (30.6) | 394 (29.9) |
| Heart failure | 828 (17.2) | 612 (17.4) | 216 (16.4) |
| Chronic kidney disease | 715 (14.8) | 558 (15.9) | 157 (11.9) |
| Imaging report available at first<br>landmark | 1,948 (40.4) | 1,473 (42.0) | 475 (36.0) |
| Patient-landmarks | 30,941 | 22,018 | 8,923 |
| Events (deterioration or death<br>within 24 h) | 2,856 (9.2) | 2,190 (9.9) | 666 (7.5) |

Values are n (%) unless stated. Patients with both ICH and SAH were coded with the reference subtype in the models; coding them in both categories changed no reported estimate (Additional file 1: Table S10).

### Imaging reports and phenotypes

Of 6,994 reports from 3,643 patients, 6,992 were processed; two could not be and were treated as unavailable (Additional file 1). An eligible report was available at 58.8% of patient-landmarks, rising from 35.6% at 6 hours to 73.4% at 48 hours, and the report used was a median of 13.0 hours old.

Three of seven candidate phenotypes met the reliability criterion — midline shift (κ 0.80), intraventricular haemorrhage (0.68) and intracranial haemorrhage (0.64) — as did four location categories: intraparenchymal (0.75) and subarachnoid (0.73) haemorrhage, middle cerebral artery territory (0.78) and cerebellum (0.66) (Table 2). Acute or subacute infarction (0.58), cerebral oedema (0.56), mass effect (0.54) and chronic ischaemic change (0.45) did not. The extracted phenotypes followed the expected clinical pattern: intracranial haemorrhage was reported at 96.0% of imaged landmarks in ICH and 96.4% in SAH but 21.2% in AIS, and middle cerebral artery territory at 37.7% in AIS but 2.1% in ICH.

**Table 2.** Agreement between automated extraction and manual annotation (200 random reports)

| Finding | $\kappa$ | Annotator-positive reports | Primary model |
| --- | --- | --- | --- |
| **Phenotypes** (four-level scale) |  |  |  |
| Midline shift | 0.80 | 42 | Yes |
| Intraventricular haemorrhage | 0.68 | 31 | Yes |
| Intracranial haemorrhage | 0.64 | 92 | Yes |
| Acute or subacute infarction | 0.58 | 52 | No |
| Cerebral oedema | 0.56 | 44 | No |
| Mass effect | 0.54 | 43 | No |
| Chronic ischaemic change | 0.45 | 39 | No |
| **Location categories** (presence) |  |  |  |
| Haemorrhage: intraparenchymal | 0.75 | 44 | Yes |
| Haemorrhage: subarachnoid | 0.73 | 31 | Yes |
| Territory: middle cerebral artery | 0.78 | 33 | Yes |
| Region: cerebellum | 0.66 | 24 | Yes |

Entry required $\kappa \geq 0.60$ and $\geq 10$ annotator-positive reports, a criterion for automated extraction rather than a claim of interchangeability with expert annotation. Reference standard: one trained annotator, blinded. Phenotypes not entering the primary model were analysed in sensitivity analysis. Herniation, large territorial infarct, hydrocephalus and large-vessel occlusion had fewer than 10 annotator-positive reports; all phenotypes and location categories are in Additional file 1: Tables S1 and S2.

### Baseline models

In validation, adding physiology and treatment to the dynamic-state baseline raised the AUROC from 0.795 to 0.849 (difference 0.054, 95% CI 0.032 to 0.077) and top-10% capture by 6.76 percentage points (2.06 to 10.82). Adding imaging availability changed neither (capture −0.15 points, −0.49 to 0.58).

### Primary comparison

Among the highest-ranked 10% of patients, the imaging-augmented model and the imaging-availability baseline captured the same number of events, 348 of 666 each: an event capture of 52.25% and a difference of 0.00 percentage points (95% CI −2.20 to +2.01) (Table 3). The interval included zero and excluded an improvement as large as the 5-point interpretation benchmark. At the 24-hour landmark, both models flagged 117 of 1,171 patients; of 84 events, the baseline captured 54.8% and the imaging-augmented model 52.4%.

**Table 3.** Model performance in temporal validation (8,923 patient-landmarks, 666 events)

|  | Dynamic-state<br>baseline (M0-state) | Full physiological<br>baseline (M0-full) | Imaging-availability<br>baseline (M0+A) | Imaging-augmented<br>(M1-D) |
| --- | --- | --- | --- | --- |
| AUROC | 0.795 | 0.849 | 0.849 | 0.855 |
| AUPRC | 0.322 | 0.391 | 0.392 | 0.399 |
| Brier score | 0.059 | 0.055 | 0.055 | 0.056 |
| Top-10% event<br>capture, % | 45.6 | 52.4 | 52.3 | 52.3 |
| Top-10% positive<br>predictive value, % | 34.1 | 39.1 | 39.0 | 39.0 |
| Top-10% lift | 4.57 | 5.24 | 5.23 | 5.23 |
| Calibration intercept | −0.21 | −0.16 | −0.16 | −0.17 |
| Calibration slope | 1.05 | 0.96 | 0.97 | 0.96 |

Primary comparison, imaging-augmented minus imaging-availability baseline: top-10% event capture 0.00 percentage points (95% CI −2.20 to +2.01).

Other measures were similar: AUROC 0.855 versus 0.849, area under the precision–recall curve 0.399 versus 0.392, and Brier score 0.0556 versus 0.0554. Both models over-predicted modestly in the later period (calibration intercept −0.17 and −0.16; mean predicted risk 8.4% in both against 7.5% observed), with calibration slopes of 0.96 and 0.97 (Figure 2); intercept-only recalibration removed the offset. Decision curves overlapped across all thresholds (Figure 3).

**Figure 1.**
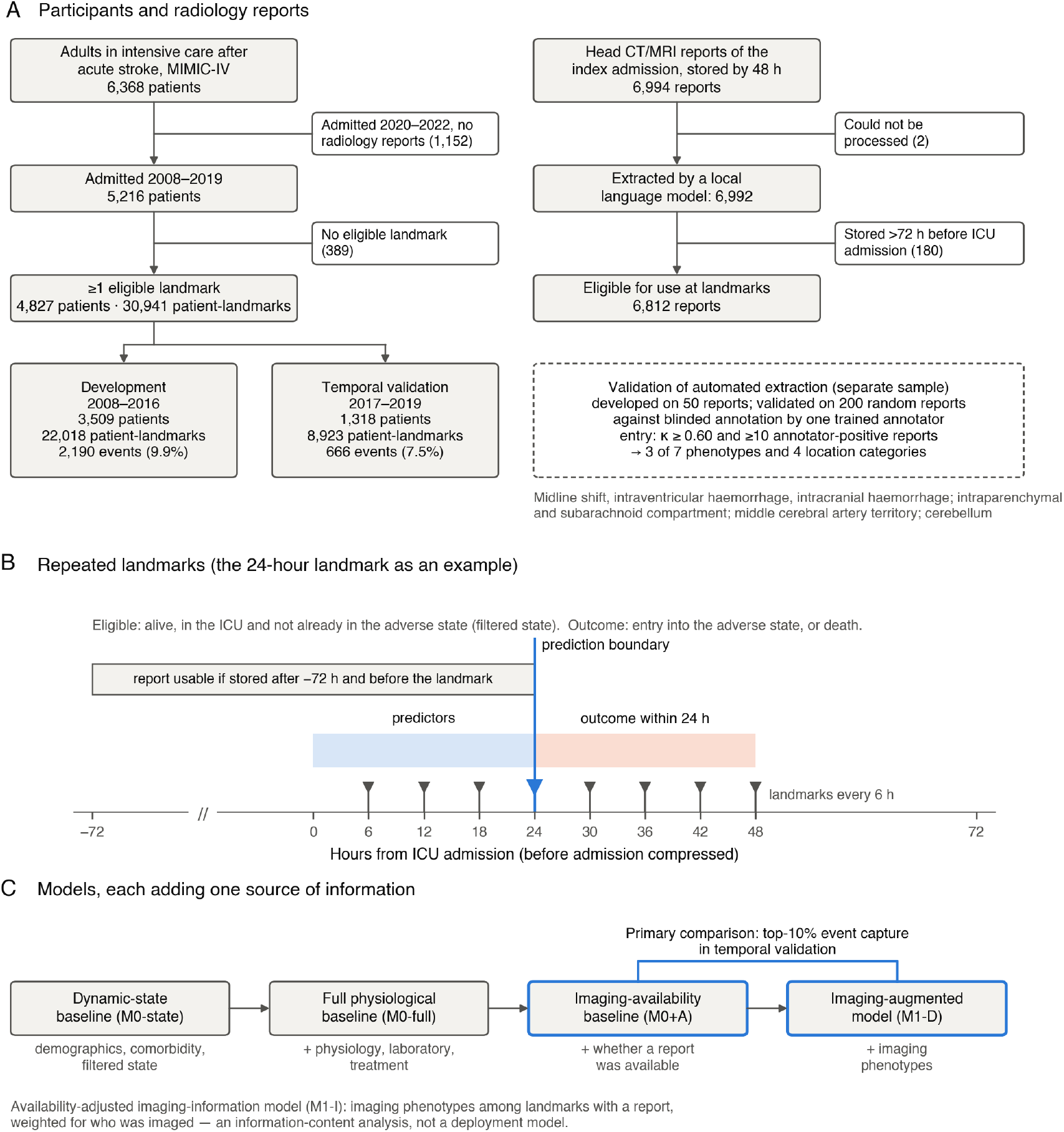
Study design. (A) Participants and radiology reports. Extraction was validated on a separate random sample of reports against blinded annotation. (B) Repeated landmarks, with the 24-hour landmark as an example: predictors use only information recorded before the landmark, including radiology reports stored no earlier than 72 hours before ICU admission, and the outcome is assessed over the following 24 hours. The hours before ICU admission are compressed. (C) Models, each adding one source of information; the primary comparison was between the imaging-availability baseline and the imaging-augmented model.

**Figure 2.**
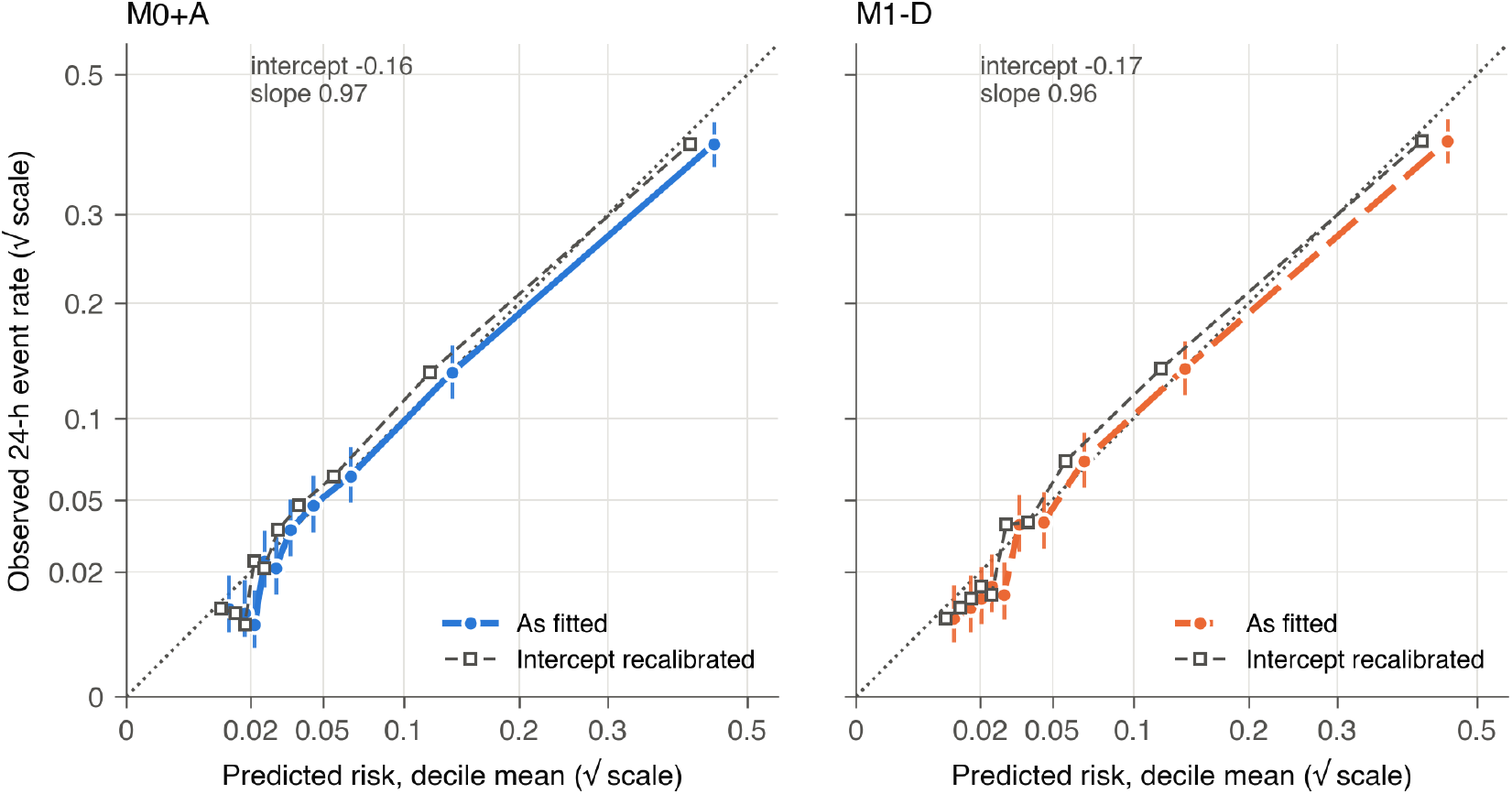
Calibration of the imaging-availability baseline and the imaging-augmented model in temporal validation. Points show observed against mean predicted 24-hour event rates by decile of predicted risk, with Wilson 95% intervals; squares show the same deciles after intercept-only recalibration. Both axes use a square-root scale; the dotted line indicates perfect calibration.

**Figure 3.**
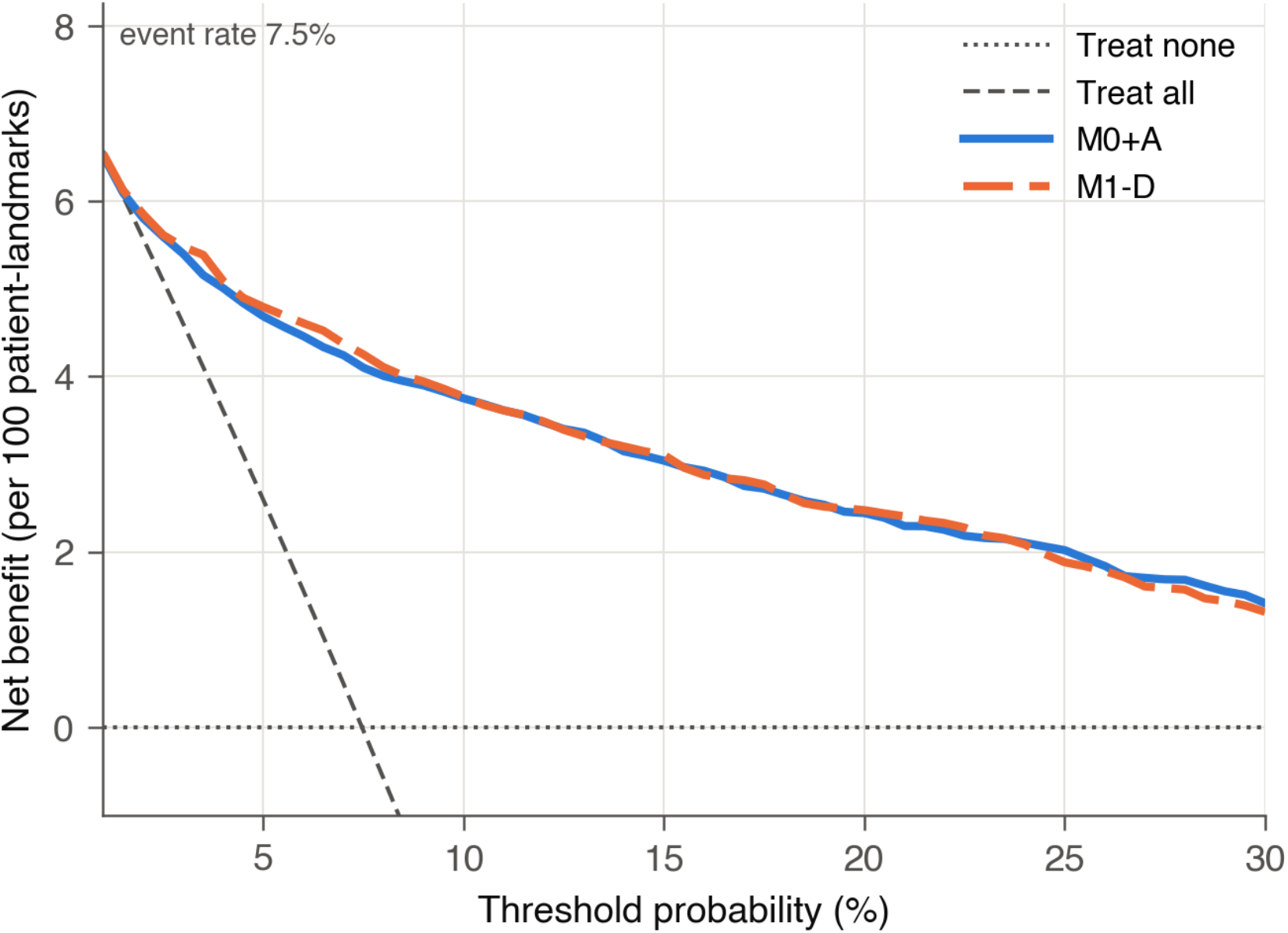
Decision curves in temporal validation. The imaging-availability baseline and the imaging-augmented model, with treat-all and treat-none strategies, for threshold probabilities of 1% to 30%. Net benefit is expressed per 100 patient-landmarks.

### Availability-adjusted imaging-information analysis (M1-I)

This analysis addressed the information content of imaging phenotypes among landmarks with an available report; it was not a deployment analysis. Among 5,127 such validation landmarks (439 events), weights were stable (0.64 to 2.25 after truncation; effective sample size 90.2% of development landmarks), and the propensity scores of imaged and non-imaged landmarks overlapped at every landmark (98.0% to 100% of imaged landmarks lay within the range of non-imaged landmarks), so no landmark was excluded. Imaging phenotypes raised the weighted AUROC from 0.820 to 0.832 but not weighted top-10% capture (−0.69 points, −4.05 to 1.45).

### Sensitivity and subtype analyses

No prespecified sensitivity analysis showed statistically supported incremental capture; point estimates ranged from −1.43 to +1.00 percentage points, and no upper confidence limit exceeded +3.53 (Table 4). Adding the four phenotypes below the reliability criterion gave −1.20 (−3.18 to 1.20). Within subtypes the difference was +0.55 (−2.09 to 2.01) in AIS, +1.80 (−3.14 to 4.10) in ICH and 0.00 (−9.33 to 3.57) in SAH, and a subtype-by-imaging interaction did not improve capture (0.00, −1.49 to 1.36).

**Table 4.** Sensitivity analyses and post-audit analyses of the primary comparison.

| Analysis | Difference in top-10% capture, percentage points (95% CI) |
| --- | --- |
| Primary analysis | 0.00 (−2.20 to 2.01) |
| *Outcome* |  |
| 12-hour horizon | −1.24 (−2.82 to 2.89) |
| Treatment-inclusive adverse state | −1.25 (−2.98 to 1.10) |
| Deterioration only, death as competing event | +0.33 (−2.31 to 2.13) |
| *Baseline and landmarks* |  |
| Without treatment predictors | −0.75 (−2.99 to 0.84) |
| Most probable state instead of state probabilities | +0.15 (−1.97 to 2.10) |
| Excluding filtered/full-sequence discordant landmarks | +1.00 (−1.24 to 2.24) |
| Excluding the 6-hour landmark | 0.00 (−2.23 to 1.93) |
| *Imaging phenotypes* |  |
| Unknown findings set to negative | 0.45 (−2.08 to 2.02) |
| Uncertain findings counted as positive | 0.00 (−1.88 to 1.99) |
| Adding phenotypes below the reliability criterion | −1.20 (−3.18 to 1.20) |
| No 72-hour limit on report age before ICU admission | +0.30 (−2.05 to 2.21) |
| 24-hour limit on report age before ICU admission | +0.15 (−2.25 to 2.17) |
| Report at most 72 hours old at the landmark | +0.15 (−2.31 to 1.91) |
| *Model class* |  |
| Gradient boosting, native handling of missing values | 0.60 (−1.52 to 3.53) |
| *Availability adjustment (M1-I)* |  |
| Availability model without physiology | −1.43 (−4.29 to 1.26) |
| Weights truncated at 2.5th and 97.5th percentiles | −0.89 (−3.87 to 1.79) |
| *Post-audit analyses (not prespecified)* |  |
| Validation patients not used to estimate the state model | +1.55 (−4.42 to 3.07) |
| Laboratory values assigned by result time | +0.30 (−2.03 to 2.02) |

### Post-audit analyses

In the two post-audit analyses, the difference in top-10% capture was +1.55 (−4.42 to 3.07) among the 405 validation patients not used to estimate the state model, and +0.30 (−2.03 to 2.02) with laboratory values assigned by result time (Table 4; Additional file 1: Tables S12 and S13). The state-only baseline did not discriminate better in the 913 patients used to estimate the state model than in the 405 who were not (AUROC 0.792 and 0.803).

### Repeated extraction with a larger model

When all reports were extracted again after the primary result was known, with a larger language model (Qwen3.8, 27 billion parameters, 4-bit quantised, run locally), the difference in top-10% capture was −0.30 percentage points (−2.70 to 1.80) with the same seven findings and −0.15 (−2.62 to 1.99) with the nine that met the reliability criterion under the new extraction (Additional file 1: Methods S6 and Table S14).

### Post hoc redundancy analysis

Imaging phenotypes were associated with subsequent deterioration: among imaged validation landmarks, the 24-hour event rate was 19.4% with midline shift and 6.2% without, and 15.4% with intraventricular haemorrhage and 6.9% without. Added to demographic and clinical characteristics with imaging availability, they raised top-10% capture from 14.4% to 27.3% (+12.91 points, 7.54 to 17.81) (Table 5, Figure 4). After the filtered state was added, the increment was no longer evident (−2.55, −5.20 to 1.51), and it remained absent after Glasgow Coma Scale scores (−0.90, −2.91 to 0.72) and then all physiology and treatment (0.00, −2.18 to 2.11) were added.

**Table 5.** Post hoc redundancy analysis: imaging phenotypes added to nested baselines.

| Baseline | AUROC, baseline<br>→ with imaging | Top-10% capture, %,<br>baseline → with imaging | Difference, percentage<br>points (95% CI) |
| --- | --- | --- | --- |
| Demographic and clinical<br>characteristics, imaging availability | 0.595 → 0.627 | 14.4 → 27.3 | +12.91 (7.54 to 17.81) |
| + filtered dynamic state | 0.801 → 0.809 | 47.3 → 44.7 | −2.55 (−5.20 to 1.51) |
| + Glasgow Coma Scale eye and<br>motor scores | 0.828 → 0.835 | 51.2 → 50.3 | −0.90 (−2.91 to 0.72) |
| + all physiology and treatment (= imaging-availability baseline) | 0.849 → 0.855 | 52.3 → 52.3 | 0.00 (−2.18 to 2.11) |

Not part of the protocol; specified in writing before it was run. Baselines are nested in the order shown, so each row adds to all rows above it; Glasgow Coma Scale scores were not examined without the dynamic state. The last row reproduces the primary comparison with a different resampling sequence.

**Figure 4.**
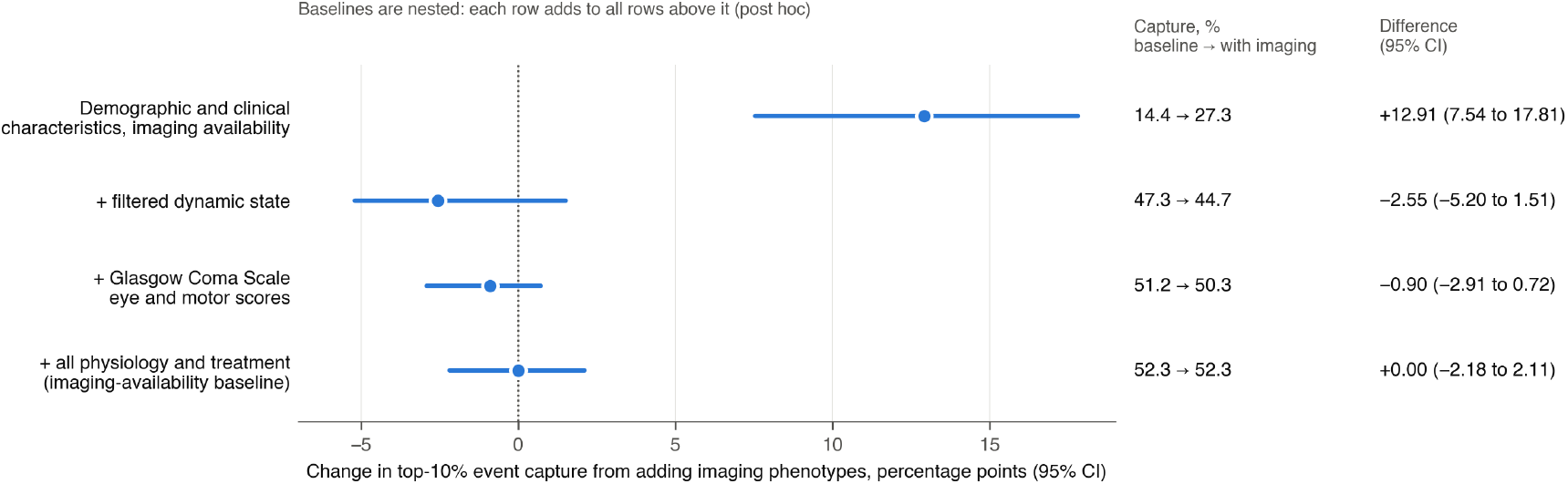
Post hoc redundancy analysis. Change in top-10% event capture in temporal validation when imaging phenotypes are added to nested baselines. Each row adds to all rows above it, so Glasgow Coma Scale scores were not examined without the dynamic state. Points show the difference and lines the 95% patient-level bootstrap confidence interval; the dotted line marks no difference.

## Discussion

This study asked a different question from most multimodal prediction work: once a model already knows how an ICU patient’s physiology is evolving after acute stroke, what do imaging phenotypes from routine radiology reports add to short-term risk stratification? We observed no added value. Among the 10% of patients ranked highest at each landmark, the phenotypes identified no additional event among 666, a difference in event capture of 0.00 percentage points (95% CI −2.20 to +2.01). The upper confidence limit, +2.01, lies below the 5-percentage-point benchmark set in advance for clinical interpretation, so the data do not support an improvement of that size.

This does not mean that the phenotypes carry no prognostic information. Added to a baseline of demographic and clinical characteristics alone, the same phenotypes raised the share of events captured in the top 10% from 14.4% to 27.3%. Once the filtered dynamic state was in the model, that gain was no longer evident. The apparent value of a new modality depends critically on the information already contained in the reference model.

One explanation consistent with these findings is that part of the lesion information recorded in routine reports overlaps with the clinical and physiological changes that the dynamic state represents. Midline shift and intraventricular haemorrhage were associated with deterioration here, but brain displacement is itself associated with depressed consciousness [4], and a lesion that is about to cause deterioration may already be affecting the bedside measurements from which the state is built. The analysis cannot identify which measurements carry this overlap. The baselines were nested, the imaging increment had already disappeared when the state was added, and consciousness scores were entered only after it, so their separate contribution was not assessed.

A second explanation cannot be excluded, and concerns how imaging was measured. This study did not evaluate CT or MRI images but a chain of representations: a scan, a radiologist’s report, automated extraction, and a small set of categories. Each step can lose information, and extraction adds its own error; only three of seven candidate phenotypes met the reliability criterion. The absence of an observed increment for report-derived phenotypes therefore cannot be read as an absence of additional predictive information in the images themselves. Quantitative imaging measures, such as lesion volume, or features derived directly from the images may carry information that these phenotypes do not.

The findings have two methodological implications. First, among imaged patients, imaging phenotypes raised the AUROC by 0.012 without improving top-10% capture. This illustrates that a modest change in global discrimination need not translate into improved case identification at a fixed clinical capacity. When a model is intended to decide which patients receive closer reassessment or monitoring, event capture at a prespecified capacity corresponds directly to that use.

Second, validating automated extraction before modelling changed which information entered the model: four clinically important phenotypes were excluded because they did not meet the reliability criterion. Adding them back did not change the result, but that does not show their measurement error to be unimportant. Setting and enforcing the criterion in advance did make the boundary between extraction error and model performance explicit.

### Limitations

Two limitations concern measurement. The study evaluated phenotypes derived from radiology reports rather than the images, and reporting, extraction and categorisation can each lose information that direct imaging features or quantitative measures might retain. Extraction was validated against a single trained annotator working blind to a written guideline, not against a multi-expert consensus; agreement between annotators was not measured, and the reliability estimates that determined which phenotypes entered the model depend on that one reference. A larger extraction model, applied after the primary result was known, agreed only slightly better with this annotator and did not change the result (Additional file 1: Table S14); it addresses the capacity of the extraction model, not the reliance on a single reference standard.

Others concern the design and generalisability. The outcome was physiological deterioration or death within 24 hours rather than functional recovery, for which imaging may matter more. Data came from one health system, and only temporal validation was possible: the dynamic states have been examined in an independent multicentre database, but that database lacks comparable radiology reports. Inverse probability weighting accounts only for measured reasons for imaging, and residual selection by unrecorded clinical judgement cannot be excluded. The models produce individual probabilities, but the evidence supports their use for ranking and prioritising groups of patients, not for individual treatment decisions. Performance was not examined across sociodemographic groups, so the fairness of the models was not assessed.

Two further limitations were inherited from the preprocessing of the earlier study and found on audit. The state model had been estimated on a random split that included most temporal-validation patients, so the temporal validation cohort was not fully independent with respect to estimation of the pre-existing state representation; the result was similar in the 405 patients not used to estimate it. Laboratory values were timed by specimen collection rather than result, so about one in six was used before its result was available, with a 90th percentile delay of 1.56 hours, whereas reports were timed by storage; reassigning laboratory values by result time did not change the result.

### Conclusions

Neuroimaging phenotypes derived from routine radiology reports were prognostic for short-term deterioration in patients in intensive care after acute stroke, but we found no evidence that they added risk stratification value beyond the dynamic ICU state and contemporaneous physiology. Post hoc analysis suggests that the information in these phenotypes may overlap substantially with that in the dynamic physiological state. Whether quantitative or image-derived information adds independent short-term predictive value over a strong dynamic clinical baseline remains to be determined.

## Additional files

**Additional file 1** (.docx). Supplementary methods and tables. Protocol and amendments; automated extraction and its validation; independent reproduction; data governance; repeated extraction with a larger model; Tables S1–S15.

**Additional file 2** (.docx). Completed TRIPOD+AI checklist, with the manuscript section in which each item is reported.

## Declarations

### Ethics approval and consent to participate

This retrospective study used de-identified data from MIMIC-IV version 3.1 and MIMIC-IV-Note version 2.2, distributed through PhysioNet under credentialed access. The collection of patient information and the creation of these research resources were reviewed by the Institutional Review Board of Beth Israel Deaconess Medical Center, which granted a waiver of informed consent and approved the data sharing initiative. Protected health information in the notes was removed before release. The corresponding author completed the CITI Program course Data or Specimens Only Research and signed the PhysioNet Credentialed Health Data Use Agreement before accessing the data. Report text was processed only on the study computer, including automated extraction by locally run language models, and was not sent to any external service. As the work involved only secondary analysis of existing de-identified data, no further institutional review board approval or informed consent was required.

### Consent for publication

Not applicable.

### Availability of data and materials

MIMIC-IV and MIMIC-IV-Note are available to credentialed users through PhysioNet and cannot be redistributed by the authors. No patient-level data or report text is shared. The analysis code, the protocol versions and the extraction prompt and rules are available at https://github.com/ping-ai685/stroke-imaging-increment-mimic; they contain no data.

### Competing interests

The authors declare that they have no competing interests.

### Funding

This research received no specific grant from any funding agency in the public, commercial, or not-for-profit sectors.

### Authors’ contributions

PL: conceptualization, data curation, formal analysis, investigation, methodology, software, validation, visualization, writing – original draft, and writing – review and editing. YX: conceptualization, methodology, formal analysis, and writing – review and editing. YZ: investigation, validation, and writing – review and editing. NR: methodology, and writing – review and editing. All authors read and approved the final manuscript.

## Supporting information

Additional file 2: TRIPOD+AI checklist

## Data Availability

https://physionet.org/content/mimiciv/3.1/

https://physionet.org/content/mimic-iv-note/2.2/

https://github.com/ping-ai685/stroke-imaging-increment-mimic

## Acknowledgements

Not applicable.

## List of abbreviations

AIS: acute ischaemic stroke
AUPRC: area under the precision–recall curve
AUROC: area under the receiver operating characteristic curve
CI: confidence interval
CT: computed tomography
ICH: intracerebral haemorrhage
ICU: intensive care unit
IQR: interquartile range
MRI: magnetic resonance imaging
SAH: subarachnoid haemorrhage
TRIPOD+AI: Transparent Reporting of a multivariable prediction model for Individual Prognosis Or Diagnosis updated for regression or machine learning methods.

## Notes

### Competing Interest Statement

The authors have declared no competing interest.

### Clinical Protocols

https://github.com/ping-ai685/stroke-imaging-increment-mimic/tree/main/08_paper3_multimodal/protocol

### Author Declarations

The Institutional Review Board of Beth Israel Deaconess Medical Center (Boston, MA, USA) waived ethical approval for this work. That Board reviewed the collection of patient information and the creation of the MIMIC-IV and MIMIC-IV-Note research resources, granted a waiver of informed consent and approved the data sharing initiative. This study is a secondary analysis of these de-identified data, accessed under the PhysioNet Credentialed Health Data Use Agreement after the corresponding author completed the required CITI Program training; no further institutional review board approval or informed consent was required.

