## Additional file 1: Supplementary methods and Tables S1-S15 for "Dynamic Risk Stratification for Short-Term Deterioration After Acute Stroke: Incremental Value of Radiology-Report-Derived Neuroimaging Phenotypes Beyond Dynamic ICU States — a retrospective cohort study with temporal validation"

### Supplementary Methods

#### S1. Protocol and amendments

The analysis protocol (version 1.2) was fixed before any validation annotation, before the extractor was fixed, and before any model containing imaging content was fitted. Amendments made before that point, and every later decision, were recorded with their date and with whether they preceded the result they governed. Decisions made after the protocol was fixed were: locking the rule for using imaging reports at landmarks, before any imaging model was fitted; adding the redundancy analysis, after the primary result was known, and therefore reported as post hoc; not seeking a second annotator; reporting the coding of patients with both ICH and SAH in this supplement rather than re-running the pipeline, decided after the corrected coding was shown to change no reported estimate (Table S10); and correcting implementation errors found during checking, none of which changed a conclusion (Table S11 and the project log); and, after an audit of the preprocessing inherited from the earlier study, two analyses with interpretation rules written down before they were run (Tables S12 and S13); and, after the primary result was known, a repetition of the extraction with a larger model, added to the protocol as version 1.3 before that model processed any report (S6, Table S14). Two further implementation errors were found after all results were known and corrected under protocol versions 1.4 and 1.5; the corrected results replace the original ones throughout (Table S15). First, the risk set had admitted landmarks at or after the patient’s recorded death, because time windows ran to ICU discharge, which for some deaths was recorded later than death; and windows after death had entered the full-sequence decoding that defines the outcome. Landmarks at or after death were removed, and windows after a death within 72 hours were removed before decoding. Second, the sensitivity analysis with a treatment-inclusive outcome state had used the wrong state of the earlier study’s model; it now uses the respiratory-support state, as specified.

#### S2. Automated extraction

Reports were processed by qwen2.5:7b (Q4_K_M quantisation) through Ollama 0.13.1 on local hardware, with temperature 0, seed 0, a context of 6,144 tokens and output constrained by a JSON schema. Deterministic post-processing verified the model output against the report text — a sentence-scoped rule for findings the report says cannot be assessed, a check that each claimed finding and location is mentioned, a rule restricting large-vessel occlusion to studies able to assess vessel patency, and cleaning of change statements — and every application was logged. The prompt, rules, schema and post-processing code were hashed into a version identifier (0347237dd3f5); every one of the 6,992 extracted records carries it. The extractor was developed on 50 reports and then fixed; it was not changed after validation.

**Reports that could not be processed.** 2 of 6,994 reports did not return a valid record within 900 seconds on either of 2 attempts. Replaying the fixed prompt showed the model repeating one anatomical region without end inside the region list; because decoding was deterministic, the failure recurred identically on every attempt. A typical record needed a median of 159 output tokens. The two reports were treated as unavailable — never as a negative finding — and were not re-run with a modified extractor.

#### S3. Validation of automated extraction

200 reports were drawn at random from the analysis population, one per patient, stratified by stroke subtype, excluding the 50 development reports and all reports of their patients. One trained annotator labelled them following a written guideline, blind to the extractor’s output. Agreement for phenotypes was Cohen’s κ on the four-level scale (present, absent, uncertain, not assessable); for location categories, κ for the presence of each category in a report. A phenotype or category entered the primary model if κ ≥ 0.60 with at least 10 annotator-positive reports (Tables S1 and S2). The criterion was an eligibility rule for automated extraction, not a claim of interchangeability with expert annotation, and no inter-rater agreement was measured.

#### S4. Independent reproduction

The primary result was recomputed by a script that imports none of the analysis code and uses different algorithms at each step: the imaging features were rebuilt with an as-of merge and cumulative maxima rather than a per-landmark loop; ranking used positional sorting; and bootstrap resampling used positional indices with a different random seed (Table S9).

#### S5. Data governance

MIMIC-IV and MIMIC-IV-Note were used under the PhysioNet credentialed data use agreement. Report text was processed only on the study computer; no report text or patient-level data was sent to an external service or shared, and none appears in this supplement.

#### S6. Repeated extraction with a larger model

After the primary result was known, the extraction was repeated with a second model to examine whether the absence of an observed increment depended on the extraction model. The analysis was added to the protocol as version 1.3 on 1 October 2026, before the second model was installed or had processed any report, together with its interpretation rule. It is reported as a post hoc analysis and does not replace the primary result.

The second model was chosen from a fixed, ordered list of candidates by technical criteria alone — valid output for four fictional reports, and a processing time compatible with the full run — without reference to agreement with any label. The first candidate met both: qwen3.8:27b (27 billion parameters, Q4_K_M quantisation), run locally through Ollama 0.35.0. The prompt, rules, output schema, generation settings and post-processing were those of the fixed extractor; the model’s reasoning output was switched off, and nothing else was changed. The second model was not developed or tuned on any report.

It was validated on the same 200 reports against the same annotator, then applied to all 6,994 reports, and the primary comparison was repeated, with the same landmark rule, models and bootstrap, on two sets of findings, namely the 7 findings of the primary model and the findings that met the reliability criterion under the new extraction — the same 7 plus cerebral oedema and the occipital region. A result was to be read as consistent with the primary result if its confidence interval included zero and its upper limit was below 5 percentage points; both were (Table S14).

111 of 6,994 reports (1.6%), 3 of them in the validation set, could not be processed: the model repeated its output until the context was full, and because decoding was deterministic the failure recurred on every attempt. They include the 2 reports that the fixed extractor could not process. As in the primary analysis, they were treated as unavailable, never as negative. Over the 6,883 reports processed by both models, agreement between them (κ) ranged from 0.68 for acute or subacute infarction to 0.91 for midline shift.

### Supplementary Tables

#### Table S1. Agreement for all annotated phenotypes (four-level scale)

| Phenotype | κ | Agreement, % | Annotator-positive reports | Use |
| --- | --- | --- | --- | --- |
| Midline shift | 0.80 | 92.5 | 42 | primary model |
| Intraventricular haemorrhage | 0.68 | 90.0 | 31 | primary model |
| Intracranial haemorrhage | 0.64 | 79.0 | 92 | primary model |
| Acute or subacute infarction | 0.58 | 81.0 | 52 | sensitivity analysis only |
| Cerebral oedema | 0.56 | 78.5 | 44 | sensitivity analysis only |
| Mass effect | 0.54 | 81.0 | 43 | sensitivity analysis only |
| Chronic ischaemic change | 0.45 | 68.5 | 39 | sensitivity analysis only |
| Herniation | 0.58 | 96.0 | 9 | not a candidate; <10 positives |
| Large territorial infarct | 0.19 | 93.0 | 8 | not a candidate; <10 positives |
| Hydrocephalus | 0.20 | 87.0 | 7 | not a candidate; <10 positives |
| Large-vessel occlusion | 0.15 | 87.5 | 3 | not a candidate; <10 positives |

200 random reports. Candidates for the primary model were the seven phenotypes listed first.

#### Table S2. Agreement for all location categories

| Field | Category | κ | Annotator-positive | Extractor-positive | Use |
| --- | --- | --- | --- | --- | --- |
| Territory | ACA | 1.00 | 3 | 3 | — |
| Territory | MCA | 0.78 | 33 | 45 | primary model |
| Territory | PCA | 0.74 | 7 | 9 | — |
| Territory | vertebrobasilar | 0.00 | 0 | 3 | — |
| Territory | watershed | 0.00 | 8 | 0 | — |
| Region | frontal | 0.46 | 30 | 71 | — |
| Region | parietal | 0.52 | 24 | 50 | — |
| Region | temporal | 0.55 | 18 | 38 | — |
| Region | occipital | 0.55 | 18 | 32 | — |
| Region | insula | 0.54 | 6 | 12 | — |
| Region | basal ganglia | 0.46 | 24 | 22 | — |
| Region | thalamus | 0.72 | 8 | 11 | — |
| Region | brainstem | 0.50 | 3 | 1 | — |
| Region | cerebellum | 0.66 | 24 | 22 | primary model |
| Haemorrhage compartment | intraparenchymal | 0.75 | 44 | 62 | primary model |
| Haemorrhage compartment | subarachnoid | 0.73 | 31 | 26 | primary model |
| Haemorrhage compartment | subdural | 0.69 | 7 | 10 | — |
| Haemorrhage compartment | epidural | — | 0 | 0 | — |

κ for the presence of each category in a report. — : not estimable (no positives on either side).

#### Table S3. Risk sets, events and imaging availability by landmark

| Landmark, h | At risk | Events (%) | Report available, % | Report age, median h | Validation at risk | Validation events |
| --- | --- | --- | --- | --- | --- | --- |
| 6 | 4,159 | 429 (10.3) | 35.6 | 6.5 | 1,170 | 96 |
| 12 | 4,282 | 410 (9.6) | 44.7 | 9.5 | 1,197 | 99 |
| 18 | 4,367 | 434 (9.9) | 54.3 | 10.9 | 1,212 | 104 |
| 24 | 4,168 | 356 (8.5) | 63.1 | 10.3 | 1,171 | 84 |
| 30 | 3,849 | 321 (8.3) | 66.4 | 13.3 | 1,110 | 74 |
| 36 | 3,581 | 323 (9.0) | 69.6 | 17.0 | 1,067 | 72 |
| 42 | 3,394 | 299 (8.8) | 72.1 | 20.2 | 1,025 | 69 |
| 48 | 3,141 | 284 (9.0) | 73.4 | 24.2 | 971 | 68 |

Both cohorts unless stated. Report age is the time from storage of the report used to the landmark.

#### Table S4. Risk stratification at the top 5%, 10% and 20%, temporal validation

| Model | Flagged | Event capture, % | Positive predictive value, % | Lift |
| --- | --- | --- | --- | --- |
| Dynamic-state baseline (M0-state) | 5% | 27.3 | 40.7 | 5.46 |
| Dynamic-state baseline (M0-state) | 10% | 45.6 | 34.1 | 4.57 |
| Dynamic-state baseline (M0-state) | 20% | 67.7 | 25.3 | 3.39 |
| Full physiological baseline (M0-full) | 5% | 33.5 | 49.9 | 6.68 |
| Full physiological baseline (M0-full) | 10% | 52.4 | 39.1 | 5.24 |
| Full physiological baseline (M0-full) | 20% | 71.0 | 26.5 | 3.55 |
| Imaging-availability baseline (M0+A) | 5% | 33.8 | 50.3 | 6.74 |
| Imaging-availability baseline (M0+A) | 10% | 52.3 | 39.0 | 5.23 |
| Imaging-availability baseline (M0+A) | 20% | 70.9 | 26.5 | 3.55 |
| Imaging-augmented model (M1-D) | 5% | 31.7 | 47.2 | 6.32 |
| Imaging-augmented model (M1-D) | 10% | 52.3 | 39.0 | 5.23 |
| Imaging-augmented model (M1-D) | 20% | 72.4 | 27.0 | 3.62 |

Ranking within each landmark, flagged patients pooled across landmarks.

#### Table S5. Availability model and overlap of propensity scores by landmark, temporal validation

| Landmark, h | n | Report available, % | Propensity, imaged | Propensity, not imaged | Imaged within non-imaged range, % |
| --- | --- | --- | --- | --- | --- |
| 6 | 1,170 | 32.3 | 0.133–0.836 | 0.115–0.804 | 99.5 |
| 12 | 1,197 | 42.1 | 0.164–0.844 | 0.160–0.871 | 100.0 |
| 18 | 1,212 | 51.6 | 0.160–0.896 | 0.189–0.897 | 99.7 |
| 24 | 1,171 | 61.2 | 0.174–0.900 | 0.244–0.871 | 99.0 |
| 30 | 1,110 | 65.0 | 0.243–0.909 | 0.284–0.883 | 98.3 |
| 36 | 1,067 | 69.2 | 0.182–0.937 | 0.365–0.904 | 98.0 |
| 42 | 1,025 | 72.1 | 0.325–0.949 | 0.350–0.929 | 98.8 |
| 48 | 971 | 72.6 | 0.506–0.958 | 0.430–0.941 | 98.4 |

Availability model AUROC 0.685. Stabilised weights truncated at the 1st and 99th percentiles ranged 0.64 to 2.25; effective sample size 11,795 of 13,073 development landmarks (90.2%). No landmark was excluded.

#### Table S6. Stroke subtypes: patients, events, imaging and phenotype prevalence

| Subtype | Cohort | Patients | Patient-landmarks | Events (%) | Report available, % |
| --- | --- | --- | --- | --- | --- |
| AIS | Development | 1,912 | 12,182 | 1,103 (9.1) | 53.5 |
| AIS | Validation | 791 | 5,438 | 366 (6.7) | 56.1 |
| ICH | Development | 1,096 | 6,555 | 740 (11.3) | 73.6 |
| ICH | Validation | 369 | 2,423 | 222 (9.2) | 66.2 |
| SAH | Development | 488 | 3,195 | 336 (10.5) | 52.8 |
| SAH | Validation | 155 | 1,038 | 78 (7.5) | 44.1 |
| ICH and SAH | Development | 13 | 86 | 11 (12.8) | 53.5 |
| ICH and SAH | Validation | 3 | 24 | 0 (0.0) | 62.5 |

| Phenotype, % positive among imaged landmarks | AIS | ICH | SAH | ICH and SAH |
| --- | --- | --- | --- | --- |
| Intracranial haemorrhage | 21.2 | 96.0 | 96.4 | 98.4 |
| Intraventricular haemorrhage | 5.2 | 41.8 | 34.1 | 63.9 |
| Midline shift | 11.3 | 34.4 | 13.7 | 33.3 |
| Compartment: intraparenchymal | 8.4 | 66.5 | 22.0 | 52.5 |
| Compartment: subarachnoid | 4.8 | 13.0 | 64.8 | 80.3 |
| Territory: MCA | 37.7 | 2.1 | 4.0 | 0.0 |
| Region: cerebellum | 17.9 | 13.4 | 3.5 | 1.6 |

Both cohorts. Prevalence among landmarks with an eligible report and a known (positive or negative) value.

#### Table S7. Primary comparison within stroke subtypes, temporal validation

| Subtype | Events | AUROC, baseline → augmented | Top-10% capture, % | Difference, pp (95% CI) |
| --- | --- | --- | --- | --- |
| AIS | 366 | 0.848 → 0.846 | 56.3 → 56.8 | +0.55 (−2.09 to 2.01) |
| ICH | 222 | 0.858 → 0.867 | 44.6 → 46.4 | +1.80 (−3.14 to 4.10) |
| SAH | 78 | 0.825 → 0.846 | 42.3 → 42.3 | 0.00 (−9.33 to 3.57) |

Ranked within landmark and subtype. Subtype-by-imaging interaction, compared with the imaging-augmented model: 0.00 (−1.49 to 1.36). Patients with both ICH and SAH had no validation events.

#### Table S8. Net benefit, temporal validation

| Threshold | Imaging-availability baseline | Imaging-augmented model | Difference (95% CI) |
| --- | --- | --- | --- |
| 5% | 4.69 | 4.79 | +0.11 (−0.03 to 0.26) |
| 10% | 3.75 | 3.77 | +0.02 (−0.12 to 0.16) |
| 15% | 3.04 | 3.11 | +0.07 (−0.06 to 0.21) |
| 20% | 2.44 | 2.48 | +0.03 (−0.09 to 0.16) |

Net benefit per 100 patient-landmarks; patient-level bootstrap confidence intervals.

#### Table S9. Independent reproduction of the primary result

| Quantity | Independent code (seed 7) | Analysis |
| --- | --- | --- |
| Imaging feature table, every cell | identical | — |
| AUROC, imaging-availability baseline | 0.849 | 0.849 |
| AUROC, imaging-augmented model | 0.855 | 0.855 |
| Difference in top-10% capture, pp | 0.000 | 0.000 |
| 95% CI | −2.14 to 1.97 | −2.20 to 2.01 |

#### Table S10. Coding of patients with both ICH and SAH

| Coding | AUROC, baseline | AUROC, augmented | Difference in top-10% capture, pp | 95% CI |
| --- | --- | --- | --- | --- |
| As analysed: reference category | 0.8490 | 0.8546 | 0.000 | −2.20 to 2.01 |
| Corrected: both ICH and SAH | 0.8488 | 0.8544 | +0.150 | −2.15 to 2.08 |

16 patients. In the models, subtype indicators were ICH and SAH, so these patients fell in the reference category with AIS.

#### Table S11. Correction to baseline-model differences quoted in the protocol

| Difference | Quoted in protocol §9.1 | Observed (reported in the manuscript) |
| --- | --- | --- |
| Full physiological minus dynamic-state baseline, AUROC | +0.055 (+0.031 to +0.077) | +0.054 (+0.032 to +0.077) |
| Imaging-availability minus full physiological baseline, AUROC | +0.001 (−0.000 to +0.001) | +0.000 (0.000 to +0.001) |
| Imaging-availability minus full physiological baseline, top-10% capture, pp | +0.01 (−0.61 to +0.56) | −0.15 (−0.49 to +0.58) |

The protocol quoted the mean of the bootstrap replicates in place of the observed difference. No conclusion changes.

#### Table S12. Temporal-validation patients used and not used to estimate the state model

| Patients | n | Events | AUROC, state-only baseline | Difference in top-10% capture, pp (95% CI) |
| --- | --- | --- | --- | --- |
| All temporal-validation patients | 1,318 | 666 | 0.795 | 0.00 (−2.20 to 2.01) |
| Not used to estimate the state model | 405 | 194 | 0.803 | +1.55 (−4.42 to 3.07) |
| Used to estimate the state model | 913 | 472 | 0.792 | 0.00 (−2.69 to 2.46) |

The state model and its preprocessing were estimated in the earlier study on a random split of its 2008–2022 cohort. The fitted models are unchanged; they are evaluated separately in the two groups. If the state model were optimistic on patients it had seen, the state-only baseline would discriminate clearly better in that group.

#### Table S13. Laboratory result times and the result-time rebuild

| Laboratory value | Values used | Resulted after the landmark, % | Delay if late, median, h | Delay if late, 90th percentile, h |
| --- | --- | --- | --- | --- |
| All eight | 101,593 | 17.4 | 0.60 | 1.56 |
| Urea nitrogen | 12,839 | 20.0 | 0.67 | 1.62 |
| Creatinine | 12,871 | 20.1 | 0.67 | 1.62 |
| Glucose | 12,692 | 20.1 | 0.67 | 1.62 |
| Haemoglobin | 11,791 | 12.1 | 0.43 | 1.31 |
| Platelets | 11,803 | 12.3 | 0.45 | 1.35 |
| Potassium | 13,660 | 20.3 | 0.67 | 1.62 |
| Sodium | 14,225 | 20.4 | 0.67 | 1.60 |
| White cells | 11,712 | 12.2 | 0.45 | 1.33 |

| Laboratory timing | AUROC, baseline | AUROC, augmented | Capture, baseline, % | Capture, augmented, % | Difference, pp (95% CI) |
| --- | --- | --- | --- | --- | --- |
| As analysed: specimen time | 0.849 | 0.855 | 52.3 | 52.3 | 0.00 (−2.20 to 2.01) |
| Rebuilt: result time | 0.847 | 0.853 | 52.4 | 52.7 | +0.30 (−2.03 to 2.02) |

Values used: the last value of each laboratory variable in each window 0–7 of the analysis patients, assigned by specimen time as in the earlier study. In the rebuild, values were assigned to the window of their result; the earlier study’s range checks, forward fill, training medians and scaling, and the fixed state model, were applied unchanged, and the laboratory predictors and filtered state recomputed. Outcome, landmarks, imaging features, models and bootstrap were those of the primary analysis. The first row, run through the same code, reproduces the primary result.

#### Table S14. Repeated extraction with a larger model

**A. Agreement with the annotator (200 random reports)**

| Finding | Annotator-positive reports | κ, fixed extractor (7B) | κ, larger model (27B) |
| --- | --- | --- | --- |
| Midline shift | 42 | 0.80 | 0.82 |
| Intraventricular haemorrhage | 31 | 0.68 | 0.71 |
| Intracranial haemorrhage | 92 | 0.64 | 0.67 |
| Cerebral oedema | 44 | 0.56 | 0.67 |
| Acute or subacute infarction | 52 | 0.58 | 0.58 |
| Chronic ischaemic change | 39 | 0.45 | 0.57 |
| Mass effect | 43 | 0.54 | 0.52 |
| Compartment: intraparenchymal | 44 | 0.75 | 0.74 |
| Compartment: subarachnoid | 31 | 0.73 | 0.84 |
| Territory: MCA | 33 | 0.78 | 0.81 |
| Region: cerebellum | 24 | 0.66 | 0.73 |
| Region: occipital | 18 | 0.55 | 0.67 |

**B. Primary comparison repeated (temporal validation, 8,923 patient-landmarks, 666 events)**

| Extraction and findings | Events captured, baseline / augmented | AUROC, baseline / augmented | Difference in top-10% capture, pp (95% CI) |
| --- | --- | --- | --- |
| Fixed extractor, 7 findings (primary result) | 348 / 348 | 0.849 / 0.855 | 0.00 (−2.20 to 2.01) |
| Larger model, the same 7 findings | 348 / 346 | 0.849 / 0.854 | −0.30 (−2.70 to 1.80) |
| Larger model, 9 findings meeting the criterion | 348 / 347 | 0.849 / 0.853 | −0.15 (−2.62 to 1.99) |

Post hoc analysis, specified in protocol version 1.3 before the larger model processed any report (Supplementary Methods S6). Panel A: four-level κ; annotator-positive counts are for all 200 reports, and the larger model’s κ is computed on the 197 it processed. Findings enter a model when κ ≥ 0.60 with at least 10 annotator-positive reports. Panel B: the baseline does not use imaging content and is the same in every row; the 9 findings are the 7 plus cerebral oedema and the occipital region. Each row was computed by the same code, which reproduces the primary result in the first row.

#### Table S15. Correction of two implementation errors found after the results were known

| Quantity | Before correction | After correction |
| --- | --- | --- |
| Development: patient-landmarks / events | 22,134 / 2,214 | 22,018 / 2,190 |
| Temporal validation: patients / patient-landmarks / events | 1,324 / 8,958 / 671 | 1,318 / 8,923 / 666 |
| AUROC, baseline / augmented | 0.850 / 0.855 | 0.849 / 0.855 |
| Primary comparison, difference in top-10% capture, pp (95% CI) | +0.15 (−2.31 to +1.91) | 0.00 (−2.20 to +2.01) |
| Treatment-inclusive outcome state: validation events; difference, pp (95% CI) | 1,081; +0.83 (−0.48 to +1.12) | 720; −1.25 (−2.98 to +1.10) |

Landmarks at or after the recorded death: 151 in the analysis period (116 development, 35 validation; 83 patients), carrying 29 events, all windows after death decoded as the adverse state. Windows after a death within 72 hours removed before decoding: 383 in 204 stays. Before correction the treatment-inclusive analysis used the neurological impairment–low support state; on the uncorrected landmarks the specified respiratory-support state gives 725 validation events. Every conclusion is unchanged.
