## Additional file 2: TRIPOD+AI checklist for "Dynamic Risk Stratification for Short-Term Deterioration After Acute Stroke: Incremental Value of Radiology-Report-Derived Neuroimaging Phenotypes Beyond Dynamic ICU States — a retrospective cohort study with temporal validation"

Checklist items reproduced from Collins GS, Moons KGM, Dhiman P, et al. TRIPOD+AI statement: updated guidance for reporting clinical prediction models that use regression or machine learning methods. BMJ 2024;385:e078378 (Table 1). D, development; E, evaluation. Locations are given by manuscript section.

| Section/topic | Item | D/E | Checklist item | Where reported |
| --- | --- | --- | --- | --- |
| Title | 1 | D;E | Identify the study as developing or evaluating the performance of a multivariable prediction model, the target population, and the outcome to be predicted | Title |
| Abstract | 2 | D;E | See TRIPOD+AI for Abstracts checklist | Abstract (structured: Background, Methods, Results, Conclusions) |
| Background | 3a | D;E | Explain the healthcare context (including whether diagnostic or prognostic) and rationale for developing or evaluating the prediction model, including references to existing models | Background, paragraphs 1–3 |
|  | 3b | D;E | Describe the target population and the intended purpose of the prediction model in the context of the care pathway, including its intended users (eg, healthcare professionals, patients, public) | Background, paragraphs 1 and 4; Discussion (ranking and prioritisation by ICU clinicians) |
|  | 3c | D;E | Describe any known health inequalities between sociodemographic groups | Not addressed; stated in Limitations |
| Objectives | 4 | D;E | Specify the study objectives, including whether the study describes the development or validation of a prediction model (or both) | Background, final paragraph |
| Data | 5a | D;E | Describe the sources of data separately for the development and evaluation datasets (eg, randomised trial, cohort, routine care or registry data), the rationale for using these data, and representativeness of the data | Methods: Data source and participants; Limitations (single health system) |
|  | 5b | D;E | Specify the dates of the collected participant data, including start and end of participant accrual; and, if applicable, end of follow-up | Methods: Data source and participants (development 2008–2016, temporal validation 2017–2019); outcome horizon 24 hours |
| Participants | 6a | D;E | Specify key elements of the study setting (eg, primary care, secondary care, general population) including the number and location of centres | Methods: Data source and participants (intensive care units of one academic centre, Boston) |
|  | 6b | D;E | Describe the eligibility criteria for study participants | Methods: Data source and participants; Landmarks, dynamic state and outcome (risk set) |
|  | 6c | D;E | Give details of any treatments received, and how they were handled during model development or evaluation, if relevant | Methods: Landmarks, dynamic state and outcome (treatment-free state); Models (treatments as predictors); Table 4 (model without treatment predictors) |
| Data preparation | 7 | D;E | Describe any data pre-processing and quality checking, including whether this was similar across relevant sociodemographic groups | Methods: Landmarks, dynamic state and outcome (timing rules, preprocessing of the earlier study); Imaging phenotypes; Additional file 1 (S2, S3, Tables S12–S13). Not examined across sociodemographic groups |
| Outcome | 8a | D;E | Clearly define the outcome that is being predicted and the time horizon, including how and when assessed, the rationale for choosing this outcome, and whether the method of outcome assessment is consistent across sociodemographic groups | Methods: Landmarks, dynamic state and outcome; Background (rationale). Assessed by the same algorithm for every patient |
|  | 8b | D;E | If outcome assessment requires subjective interpretation, describe the qualifications and demographic characteristics of the outcome assessors | Not applicable: the outcome is derived algorithmically from the state model and recorded death times |
|  | 8c | D;E | Report any actions to blind assessment of the outcome to be predicted | Not applicable: algorithmic outcome; the state model was fixed before this study and used no outcome data (Methods) |
| Predictors | 9a | D | Describe the choice of initial predictors (eg, literature, previous models, all available predictors) and any pre-selection of predictors before model building | Methods: Models (predictors fixed in the protocol, from the earlier study); Imaging phenotypes (reliability criterion) |
|  | 9b | D;E | Clearly define all predictors, including how and when they were measured (and any actions to blind assessment of predictors for the outcome and other predictors) | Methods: Models; Landmarks, dynamic state and outcome (timing); Imaging phenotypes (storage time, update rule) |
|  | 9c | D;E | If predictor measurement requires subjective interpretation, describe the qualifications and demographic characteristics of the predictor assessors | Methods: Imaging phenotypes (automated extraction; validation against one trained annotator, blinded to extractor output); Limitations |
| Sample size | 10 | D;E | Explain how the study size was arrived at (separately for development and evaluation), and justify that the study size was sufficient to answer the research question. Include details of any sample size calculation | Methods: Statistical analysis (all eligible patients; pooling across landmarks for precision; minimum events for the subtype interaction). No formal sample size calculation |
| Missing data | 11 | D;E | Describe how missing data were handled. Provide reasons for omitting any data | Methods: Landmarks, dynamic state and outcome (imputation from the earlier study); Imaging phenotypes (unknown findings coded separately); Table 4 (native missing-value handling) |
| Analytical methods | 12a | D | Describe how the data were used (eg, for development and evaluation of model performance) in the analysis, including whether the data were partitioned, considering any sample size requirements | Methods: Data source and participants (temporal split); Models (fitted in development, applied unchanged to validation) |
|  | 12b | D | Depending on the type of model, describe how predictors were handled in the analyses (functional form, rescaling, transformation, or any standardisation) | Methods: Models (standardised predictors); Landmarks, dynamic state and outcome (state probabilities as log-ratios and entropy) |
|  | 12c | D | Specify the type of model, rationale, all model building steps, including any hyperparameter tuning, and method for internal validation | Methods: Models (L2-penalised logistic regression, stepwise ladder, fixed penalty, no tuning); evaluation by temporal validation |
|  | 12d | D;E | Describe if and how any heterogeneity in estimates of model parameter values and model performance was handled and quantified across clusters (eg, hospitals, countries). See TRIPOD-Cluster for additional considerations | Not applicable: one centre. Patient-level clustering of landmarks handled in the bootstrap (Methods: Statistical analysis) |
|  | 12e | D;E | Specify all measures and plots used (and their rationale) to evaluate model performance (eg, discrimination, calibration, clinical utility) and, if relevant, to compare multiple models | Methods: Statistical analysis (primary comparison, secondary analyses) |
|  | 12f | E | Describe any model updating (eg, recalibration) arising from the model evaluation, either overall or for particular sociodemographic groups or settings | Methods: Statistical analysis (intercept-only recalibration) |
|  | 12g | E | For model evaluation, describe how the model predictions were calculated (eg, formula, code, object, application programming interface) | Declarations: Availability of data and materials (analysis code) |
| Class imbalance | 13 | D;E | If class imbalance methods were used, state why and how this was done, and any subsequent methods to recalibrate the model or the model predictions | Not applicable: no class imbalance methods were used |
| Fairness | 14 | D;E | Describe any approaches that were used to address model fairness and their rationale | Not addressed; stated in Limitations |
| Model output | 15 | D | Specify the output of the prediction model (eg, probabilities, classification). Provide details and rationale for any classification and how the thresholds were identified | Methods: Statistical analysis (predicted probabilities ranked within landmark; top 10% as a fixed capacity, with 5% and 20%) |
| Training versus evaluation | 16 | D;E | Identify any differences between the development and evaluation data in healthcare setting, eligibility criteria, outcome, and predictors | Methods: Data source and participants; Results: Participants and Baseline models (fall in event rate); Table 1 |
| Ethical approval | 17 | D;E | Name the institutional research board or ethics committee that approved the study and describe the participant informed consent or the ethics committee waiver of informed consent | Methods: Data source and participants; Declarations: Ethics approval and consent to participate |
| Funding | 18a | D;E | Give the source of funding and the role of the funders for the present study | Declarations: Funding |
| Conflicts of interest | 18b | D;E | Declare any conflicts of interest and financial disclosures for all authors | Declarations: Competing interests |
| Protocol | 18c | D;E | Indicate where the study protocol can be accessed or state that a protocol was not prepared | Methods: Study design; Additional file 1 (S1); protocol versions in the code repository |
| Registration | 18d | D;E | Provide registration information for the study, including register name and registration number, or state that the study was not registered | Methods: Study design (not registered) |
| Data sharing | 18e | D;E | Provide details of the availability of the study data | Declarations: Availability of data and materials |
| Code sharing | 18f | D;E | Provide details of the availability of the analytical code | Declarations: Availability of data and materials |
| Patient and public involvement | 19 | D;E | Provide details of any patient and public involvement during the design, conduct, reporting, interpretation, or dissemination of the study or state no involvement | Methods: Study design (no involvement) |
| Participants | 20a | D;E | Describe the flow of participants through the study, including the number of participants with and without the outcome and, if applicable, a summary of the follow-up time. A diagram may be helpful | Results: Participants; Figure 1 |
|  | 20b | D;E | Report the characteristics overall and, where applicable, for each data source or setting, including the key dates, key predictors (including demographics), treatments received, sample size, number of outcome events, follow-up time, and amount of missing data. A table may be helpful. Report any differences across key demographic groups | Results: Participants; Table 1; Additional file 1 (Tables S3, S6) |
|  | 20c | E | For model evaluation, show a comparison with the development data of the distribution of important predictors (demographics, predictors, and outcome) | Table 1 (development and temporal validation columns) |
| Model development | 21 | D;E | Specify the number of participants and outcome events in each analysis (eg, for model development, hyperparameter tuning, model evaluation) | Results: Participants; Tables 3 and 4 |
| Model specification | 22 | D | Provide details of the full prediction model (eg, formula, code, object, application programming interface) to allow predictions in new individuals and to enable third party evaluation and implementation, including any restrictions to access or reuse (eg, freely available, proprietary) | Methods: Models (predictors and model form); analysis code (Declarations). Coefficients are not reported: the models were built to compare information sources, not for deployment |
| Model performance | 23a | D;E | Report model performance estimates with confidence intervals, including for any key subgroups (eg, sociodemographic). Consider plots to aid presentation | Results; Table 3; Figures 2–3; Additional file 1 (Tables S4, S7, S8) |
|  | 23b | D;E | If examined, report results of any heterogeneity in model performance across clusters. See TRIPOD-Cluster for additional details | Not applicable: one centre |
| Model updating | 24 | E | Report the results from any model updating, including the updated model and subsequent performance | Results: Baseline models / calibration (intercept-only recalibration removed the offset); Figure 2 |
| Interpretation | 25 | D;E | Give an overall interpretation of the main results, including issues of fairness in the context of the objectives and previous studies | Discussion. Fairness not examined (Limitations) |
| Limitations | 26 | D;E | Discuss any limitations of the study (such as a non-representative sample, sample size, overfitting, missing data) and their effects on any biases, statistical uncertainty, and generalisability | Discussion: Limitations |
| Usability of the model in the context of current care | 27a | D | Describe how poor quality or unavailable input data (eg, predictor values) should be assessed and handled when implementing the prediction model | Methods: Imaging phenotypes (unavailable or unprocessable reports treated as unavailable). Implementation not addressed: the study evaluates added value, not a model for deployment |
|  | 27b | D | Specify whether users will be required to interact in the handling of the input data or use of the model, and what level of expertise is required of users | Not applicable: not a model for deployment |
|  | 27c | D;E | Discuss any next steps for future research, with a specific view to applicability and generalisability of the model | Discussion; Limitations |
